# Accuracy of machine learning-based prediction of medication adherence in clinical research

**DOI:** 10.1101/2020.05.27.20113597

**Authors:** Vidya Koesmahargyo, Anzar Abbas, Li Zhang, Lei Guan, Shaolei Feng, Vijay Yadav, Isaac R. Galatzer-Levy

## Abstract

**Background:** Medication non-adherence represents a significant barrier to treatment efficacy. Remote, real-time measurement of medication dosing can facilitate dynamic prediction of risk for medication non-adherence, which in-turn allows for proactive clinical intervention to optimize health outcomes. We examine the accuracy of dynamic prediction of non-adherence using data from remote real-time measurements of medication dosing.

**Methods:** Participants across a large set of clinical trials (*n* = 4,182) were observed via a smartphone application that video records patients taking their prescribed medication. The patients’ primary diagnosis, demographics, and prior indication of observed adherence/non-adherence were utilized to predict (1) adherence rates ≥ 80% across the clinical trial, (2) adherence ≥ 80% for the subsequent week, and (3) adherence the subsequent day using machine learning-based classification models.

**Results:** Empirically observed adherence was demonstrated to be the strongest predictor of future adherence/non-adherence. Collectively, the classification models accurately predicted adherence across the trial (AUC = 0.83), the subsequent week (AUC = 0.87) and the subsequent day (AUC = 0.87).

**Conclusions:** Real-time measurement of dosing can be utilized to dynamically predict medication adherence with high accuracy.

## Introduction

Prediction of medication adherence has traditionally relied on static and group-based factors like medication tolerability, diagnosis, treatment length, and demographics.^1,2^ With the development of technologies that automate the real-time measurement of medication dosing,^3-6^ there is an opportunity to utilize continuous data sources to dynamically predict medication adherence and proactively intervene if needed.

Adherence can be impacted by multiple factors. This can include logistical reasons such as simple forgetfulness, complexity of regimen, complications with prescription refills, side effects, adverse effects, or lack of insurance coverage for a medication.^7-9^ Adherence can also be impacted by the patient’s psychiatric/neurological status if the patient is experiencing hopelessness/helplessness (e.g. depression),^10,11^ lack of insight (e.g. schizophrenia),^12^ or behavioral avoidance (e.g. mood & anxiety disorders)^13^, and cognitive decline(e.g. Alzheimer’s, traumatic brain injury).

Many patients lack the access to care necessary to address factors affecting medication adherence as they emerge. Often, small bouts of non-adherence lead to long droughts in medication adherence. As a consequence, patients experience increases in symptom severity and treatment failure leading to hospitalizations and escalation of care.^12,13^ Clinicians need simple and straightforward ways to identify non-adherence and intervene in order to help patients stay on track of treatment regimens. Assistive technology can be effective in allowing hospitals to better focus their resources.

A number of technologies have emerged to directly track patient adherence by remotely measuring dose dosing. Methods include the use of pills that broadcast a signal when ingested^14^ and technologies that remotely observe patients ingesting medication through video on their smartphone.^15^ The data sources collected provide continuous measurement of each dosing event. Such data sources allow for dynamic and individual-level risk prediction rather than static population-level prediction. Dynamic prediction of risk can allow for efficient deployment of known effective treatment resources. Optimizing for efficacy in the context of adherence is necessary due to the widespread and diverse nature of adherence issues in clinical treatment and limited resources to manage patient care.^16^

## Methods

Here, we test the accuracy of medication dosing data to predict medication non-adherence. We built and tested algorithms to predict adherence in the context of clinical research and population health. First, a cut-off for acceptable adherence was set at 80%, consistent with clinical standards^17^. Then, we tested the predictive accuracy of static (demographic, diagnosis, treatment length, ect.) and dynamic (dosing, clinic interventions, etc.) features. We examined the predictive accuracy across a study based on early dosing behavior (first week and first two weeks of dosing) and real-time prediction during a study (prediction of adherence next week, next day). These scenarios were designed to support decision-making in the context of clinical trials, where researchers have the option to remove study participants early on in a trial (first one or two weeks) if they demonstrate non-adherence during a lead-in period.^18^ These same models can also be utilized by clinicians to estimate risk of non-adherence in outpatient therapy.

### Data collection

#### Software platform

For all analyses, data collected by the AiCure software platform (www.aicure.com) was used.^5,19^ The smartphone-based platform uses computer vision technology for confirmation of medication adherence in clinical trials, allowing for real-time adherence monitoring. It can replicate or supplement directly observed therapy (DOT). The software includes a patient-facing mobile application, which sets reminders for prescribed medications per treatment regimen. At the designated time, the participant is prompted to follow an automated dosing protocol. The video of the dosing captured through the smartphone front-facing camera is used to confirm medication ingestion using automated computer vision-based methods. This individual dose-level information is made accessible in real time to the clinician and site coordinators of the study using a clinician-facing web application.

Human review of dosing videos provides additional verification to augment the computer vision-based measurement of adherence. The purpose of this review process includes verification/confirmation of medication dosing and labelling of any intentional non-adherent/deceptive behavior. Videos of medication not being taken as instructed, or where suspicious behavior was found during video review of the medication’s intake, are labelled with a set of flags for downstream adjustment of adherence.

In addition to data on medication adherence, other study-related information was also collected, outlined in the predictive features section. The participants in this dataset consists of patients enrolled in clinical trials that have used the software platform between 2012 and 2019. Study participants with less than a week of data and studies with missing trial information were excluded.

#### Definition of adherence

The software’s measure of adherence is a binary indicator of whether or not a patient took all prescribed medications during a given dosing event. Patients can have either one or several dosing events in a day. For all analyses conducted, medication adherence was consolidated by day. Hence, adherence was defined as follows:

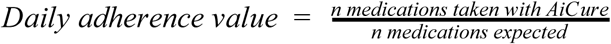

Additionally, human review included observations of non-adherent or deceptive behavior. This was used to adjust the daily adherence calculation. These observations were recorded through the use of ‘red’ or ‘orange’ alerts. A ‘red’ alert indicated strong visual evidence of intentional non-adherence, while an ‘orange’ alert indicated the presence of suspicious behavior but no visual confirmation of intentional non-adherence. The daily adherence value was then adjusted using these alerts:

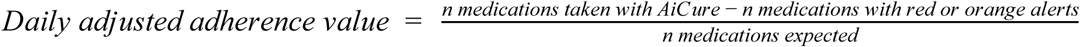

This definition of adherence was used to build classification algorithms for prediction of medication adherence above or below an 80% threshold. The ‘adherent’ class (class 0) is defined as an individual with an average adherence of 80% or higher, while the ‘non-adherent’ (class 1) is defined as an individual with an average adherence below 80%.

### Predictive features

In addition to medication adherence itself, additional study-specific characteristics were used as predictive features in the classification algorithms, outlined in this section.

#### Condition

Condition refers to the patient’s clinical condition or disease. This is indicated by the therapeutic area (e.g. schizophrenia, major depressive disorder, HIV, etc.) of the study. In cases where the therapeutic area of multiple studies share similar pathologies, these conditions are grouped together under the broader definition as shown in Table 1. For example, opioid dependence and alcohol use disorder were both placed in the category of addiction.

**Table 1:** Demographic information of participants.

| Item | Participants<br>*n* = 4,182 |
| --- | --- |
| Age, mean ± standard deviation | 39.0 ± 11.7 |
| Sex, *n* (%) |  |
| Male | 1,006 (24.1%) |
| Female | 555 (13.3%) |
| Unknown | 2,621 (62.6%) |
| Race, *n* (%) |  |
| White | 461 (11.0) |
| Black or African American | 195 (4.67) |
| Multiracial | 25 (0.60) |
| Asian | 24 (0.57) |
| Other | 22 (0.53) |
| Latino | 15 (0.36) |
| American Indian or Alaskan Native | 4 (0.10) |
| Native Hawaiian or other Pacific Islander | 4 (0.10) |
| Unknown | 3432 (82.1) |
| **Condition, *n* (%)** |  |
| **Schizophrenia** | **970 (23.3%)** |
| Attenuated psychosis syndrome | 36 (0.86%) |
| Cognitive impairment in schizophrenia | 538 (12.9%) |
| Schizophrenia or schizoaffective disorder and negative symptoms | 34 (0.81%) |
| Unspecified | 362 (8.7%) |
| **Major depressive disorder** | **777 (18.6%)** |
| Major depressive disorder with anxious distress | 20 (0.48%) |
| Depression and acute suicidal ideation behavior | 21 (0.50%) |
| Unspecified | 736 (17.6%) |
| **Addiction** | **632 (15.1%)** |
| Opioid dependence | 25 (0.60%) |
| Opioid use disorder | 5 (0.12%) |
| Alcohol use disorder | 25 (0.60%) |
| Unspecified | 577 (13.8%) |
| **HIV** | **86 (2.06%)** |
| **Bipolar depressive disorder** | **27 (0.65%)** |
| **Attention-deficit hyperactivity disorder** | **1115 (26.7%)** |
| **Healthy volunteers** | **340 (8.13%)** |
| **Post-traumatic stress disorder** | **86 (2.06%)** |
| **Hepatitis C** | **68 (1.63%)** |
| **Congestive Heart Failure** | **39 (0.93%)** |
| **Psoriasis** | **32 (0.77%)** |
| **Epilepsy** | **7 (0.17%)** |
| **Parkinson's Disease** | **2 (0.05%)** |
| **Chronic Obstructive Pulmonary Disease** | **1 (0.02%)** |

#### Interventions

Interventions refer to efforts made by study coordinators to contact patients to address any non-adherence. Study coordinators and clinicians are able to continuously monitor the participants’ course of treatment and may choose to intervene in response to patients demonstrating low or non-adherence. As the reasons for non-adherence may range from simple forgetfulness to intentional non-adherence and adverse effects, interventions help resolve these issues. Clinicians can intervene through texts, emails, phone calls, and in-person visits. The type of intervention could contain valuable information as different modes may indicate different levels of escalation. The number of interventions received for each day a patient is enrolled in a trial, both by type and in total, was used as a feature. Since data is only available when there is an occurance of an intervention, we set the assumption that days with no records indicate 0 interventions as a dichotomous yes/no item.

#### Micro-reimbursements

Micro-reimbursements incentivize patients to continue using the platform to report medication adherence by providing monetary benefit. Micro-reimbursement status is a binary indicator on whether the particular study offered micro-reimbursements to patients for reporting adherence through the software platform.

#### Trial length

Trial length is the duration in weeks in which the treatment is expected to be completed. In cases where the expected trial length can vary, the average length is used as an estimate.

#### Dose Delay

Dose delay is the length of time in minutes for a patient to begin a dosing event on the smartphone application after an alarm has prompted them to do so. The delay is only calculated for completed dosing events. If the patient starts and completes the event before the alarm time, the dose delay is a negative value. The daily average values for dose delay are then quantized to prevent overfitting when using continuous values with a broad range. The intervals used were < 30 minutes, < 2 hours, < 4 hours, < 8 hours, and > 8 hours.

#### Dose length

Dose length is the length of time in seconds it took for the patient to complete a dosing event. Longer durations may reflect difficulty in completing the medication dosing protocol and affect future behavior. Daily averages for dose length are quantized similarly to ‘dose delay’ to create the following intervals: < 30 seconds, < 2 minutes, < 5 minutes, < 10 minutes, and > 10 minutes.

### Model development

The classification of adherence was divided into four distinct models to predict a target outcome at different time points within a clinical trial. The daily values for adherence, adjusted adherence, number of interventions, dose delay and dose length were used as dynamic features at varying intervals depending on the model. The remaining features (condition, trial length, micro reimbursements) served as static predictors in all model types. Sub-datasets consisting of common features and daily features were created based on the intervals specified.

The different classification types are as follows:

1. Predicting remaining trial adherence based on the first week of the trial
2. Predicting remaining trial adherence based on the first two weeks of the trial
3. Predicting next week adherence based on the previous week in the trial
4. Predicting next day adherence based on the previous week in the trial

The classifier used for all model types was XGBoost (Extreme Gradient Boosting), which is a class of decision tree-based machine learning algorithms. The use of this algorithm was informed by a benchmark study testing for fast computation time and good model performance given structured data frames compared to other decision tree-based methods^20^. A decision tree represents the possible ways to reach a decision based on certain criteria. Random forest is a popular machine learning algorithm that uses an ensemble of decision trees, but chooses a random subset of features as its criteria. Instead of creating decision trees at random, trees can be built sequentially to minimize errors from previous models while ‘boosting’ higher performing models using a gradient descent algorithm.^21^ XGBoost was developed by combining this ensemble of techniques with even more added optimizations. With XGBoost, decision trees are pruned based on a max depth parameter instead of continuously splitting nodes to improve computational performance and avoid overfitting.^22^

Splitting of the data was done at the patient level, to ensure that no samples used during model training and model validation came from the same patient. Each model was assessed through 5-fold cross validation instead of a single split between training and test sets. This method split the dataset into 5 equal folds, each fold used as a holdout set to test the model trained on the remaining 4 folds, essentially creating a model trained on 80% of patients and tested on 20% of patients. This removes the assumption that the data split during training and testing share a similar distribution, and the model’s performance was evaluated for each fold. The performance metrics used to evaluate the classification model are detailed below, where TP: True Positives; TN: True Negatives; FP: False Positives; FN: False Negatives.

**Accuracy** = TP + TN / (TP + TN + FP + FN)

**Precision** = TP/ (TP + FP)

**Recall** = TP / (TP + FN)

**True Positive Rate** = TP/ (TP + FN)

**False Positive Rate** = FP / (FP + TN)

**ROC and AUC**. The Receiver Operating Characteristic (ROC) Curve shows the relationship between the true positive rate and false positive rate at certain thresholds of a binary classifier.

The performance of the classifier was summarized by the area under the ROC curve (AUC). With 5-fold cross validation, the AUC was calculated for each repeated iteration, and the average AUC was reported as the measure of classification performance. A grid search with cross validation was utilized to optimize the model, though only a few parameters in a small parameter space were tested (number of trees or estimators, maximum tree depth, and minimum child weight, learning rate; three different values for each parameter) to minimize search time.

## Results

### Data collected

The demographic and clinical characteristics of the patient cohort in the dataset are outlined in Table 1. The large proportion of missing values in demographic characteristics led to their exclusion as predictive features. Figure 1 (top) shows that the majority of patients in the dataset were adherent at a threshold of 80%, while Figure 1 (top) shows the breakdown of interventions by type, which was calculated by averaging daily adherence over the course of the trial at the individual level.

**Figure 1:**
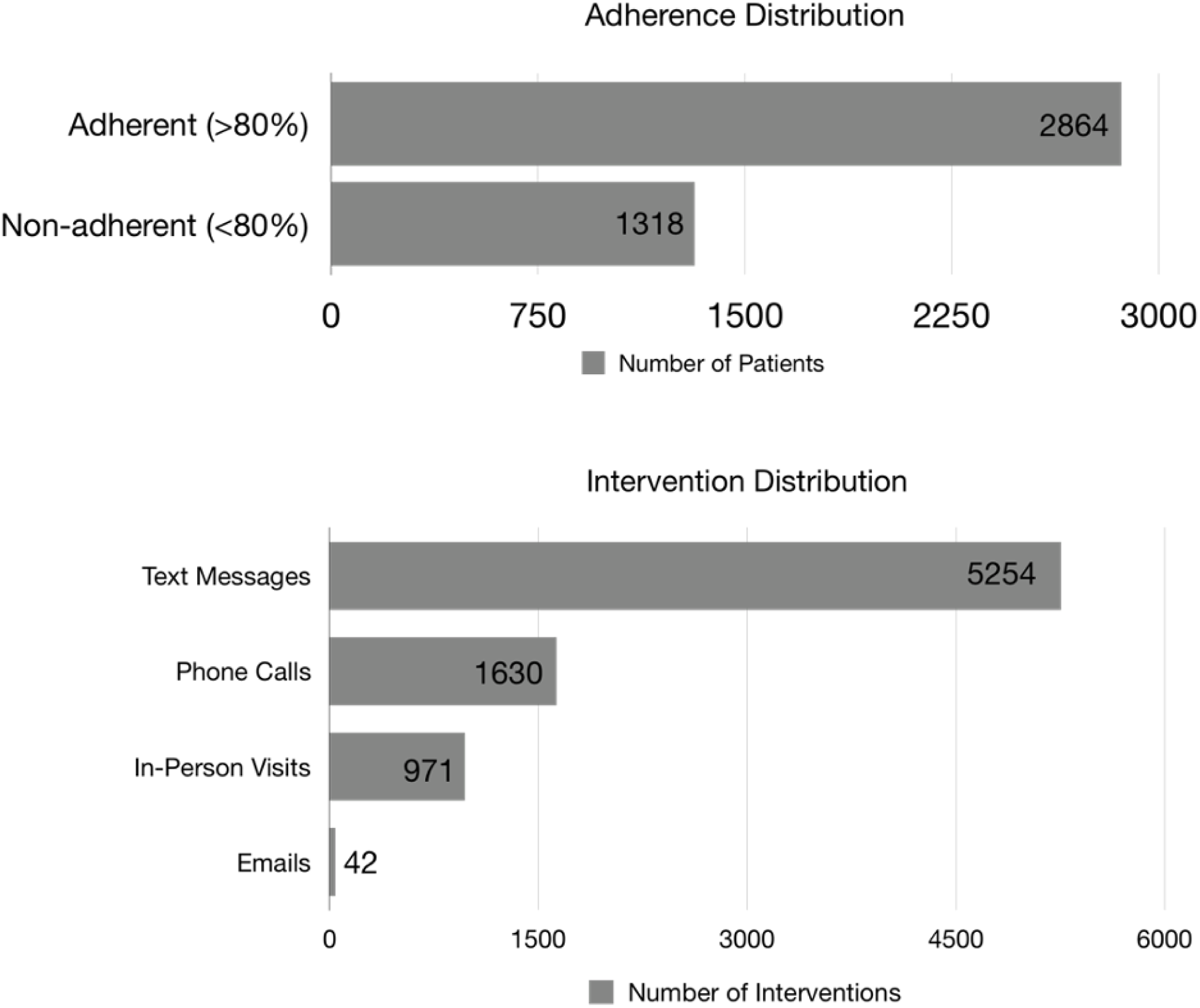
Distribution of medication adherence across all participants (top) and distribution of intervention types used (bottom).

### Predictive models

The models performed well with comparable values across the different prediction types. Performance metrics values were averaged across the five folds in Table 2. The holdout set sizes used to test the models trained on the first week and first two weeks of behavior were 863 and683 samples respectively. In contrast with the holdout set when testing models trained on previous week behavior, which were 6,623 samples for next week prediction and 45,312 samples for next day prediction. The increase in model performance was likely due to this increase in sample size.

**Table 2:**
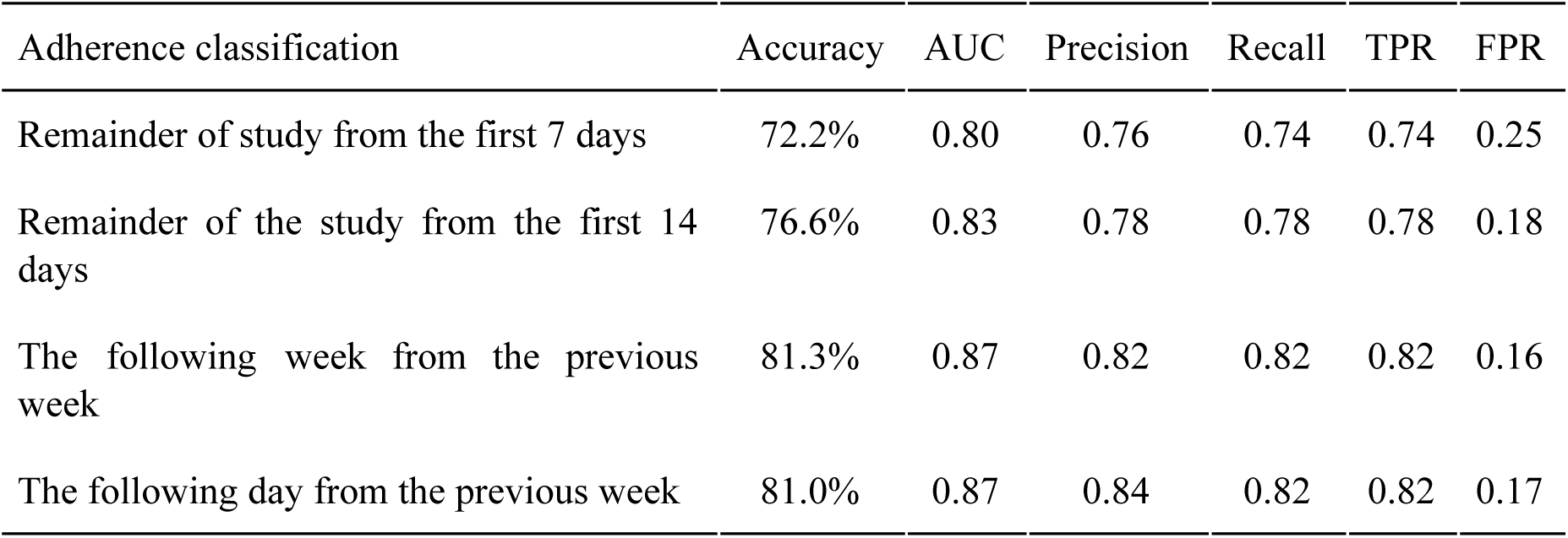
Results from the four classification types averaged across 5-fold cross validation. TPR: True Positive Rate; FPR: False Positive Rate

| Adherence classification | Accuracy | AUC | Precision | Recall | TPR | FPR |
| --- | --- | --- | --- | --- | --- | --- |
| Remainder of study from the first 7 days | 72.2% | 0.80 | 0.76 | 0.74 | 0.74 | 0.25 |
| Remainder of the study from the first 14 days | 76.6% | 0.83 | 0.78 | 0.78 | 0.78 | 0.18 |
| The following week from the previous week | 81.3% | 0.87 | 0.82 | 0.82 | 0.82 | 0.16 |
| The following day from the previous week | 81.0% | 0.87 | 0.84 | 0.82 | 0.82 | 0.17 |

Since the parameters tested during grid search did not contribute a significant increase in performance during cross validation, the results presented in Table 2 and Figure 2 were achieved using the same set of hyperparameters for the four different model types. With the exception of the number of trees (n_estimators = 550), other parameters that differed from the classifier’s default were chosen to discourage overfitting: learning rate of 0.01, maximum tree depth of 5, and minimum child weight of 6.

**Figure 2:**
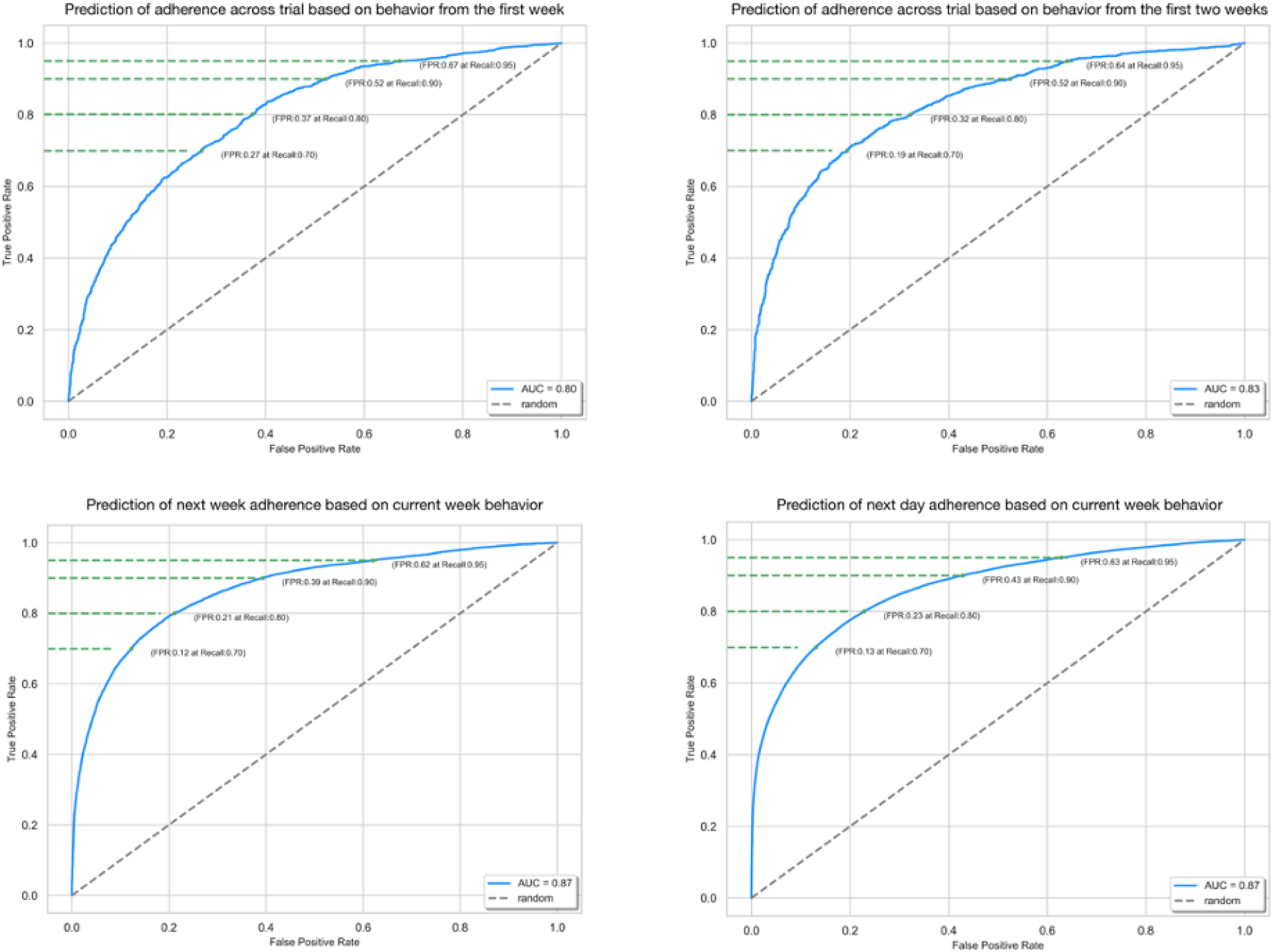
ROC curves with mean AUCs from cross-validation with specific recall thresholds of the four different model types. **(a)** Prediction of adherence across the trial based on the first week **(b)** Prediction of adherence across the trial based on the first two weeks **(c)** Prediction of next week adherence based on current week **(d)** Prediction of next day adherence based on current week.

Figure 2 shows the Receiver Operating Curves (ROCs) of the four different model types with comparable results. The model using the first two weeks of behavior performed better than the model using only the first week, and the AUC in the next week prediction is slightly better than the AUC in the next day prediction. Due to the rounding of the AUC values, the two validation results are effectively the same, but Figure 2 illustrates the slightly lower false positive rates in the next week prediction at the same recall value as the next day prediction. When predicting the next week’s average adherence, the additional data points help normalize the mean, whereas prediction of adherence is more likely to fluctuate from one day to the next.

To investigate the contribution of individual predictive features in the different adherence model types, we retrieved the feature importance from each of the cross validation folds and aggregated the results as seen in Figure 3. Feature importance was measured in terms of the average gain in accuracy across all splits when using a particular feature. Average adherence (avg_adh) has the highest relative importance regardless of adherence model type, and average adherence and condition are consistently included in each model type’s highest importance predictors. Features with considerable importance also include the average adherence values in the last few days before prediction (avg_day7, avg_day6, avg_day13), and the adjusted average adherence (adh_adj_avg).

**Figure 3:**
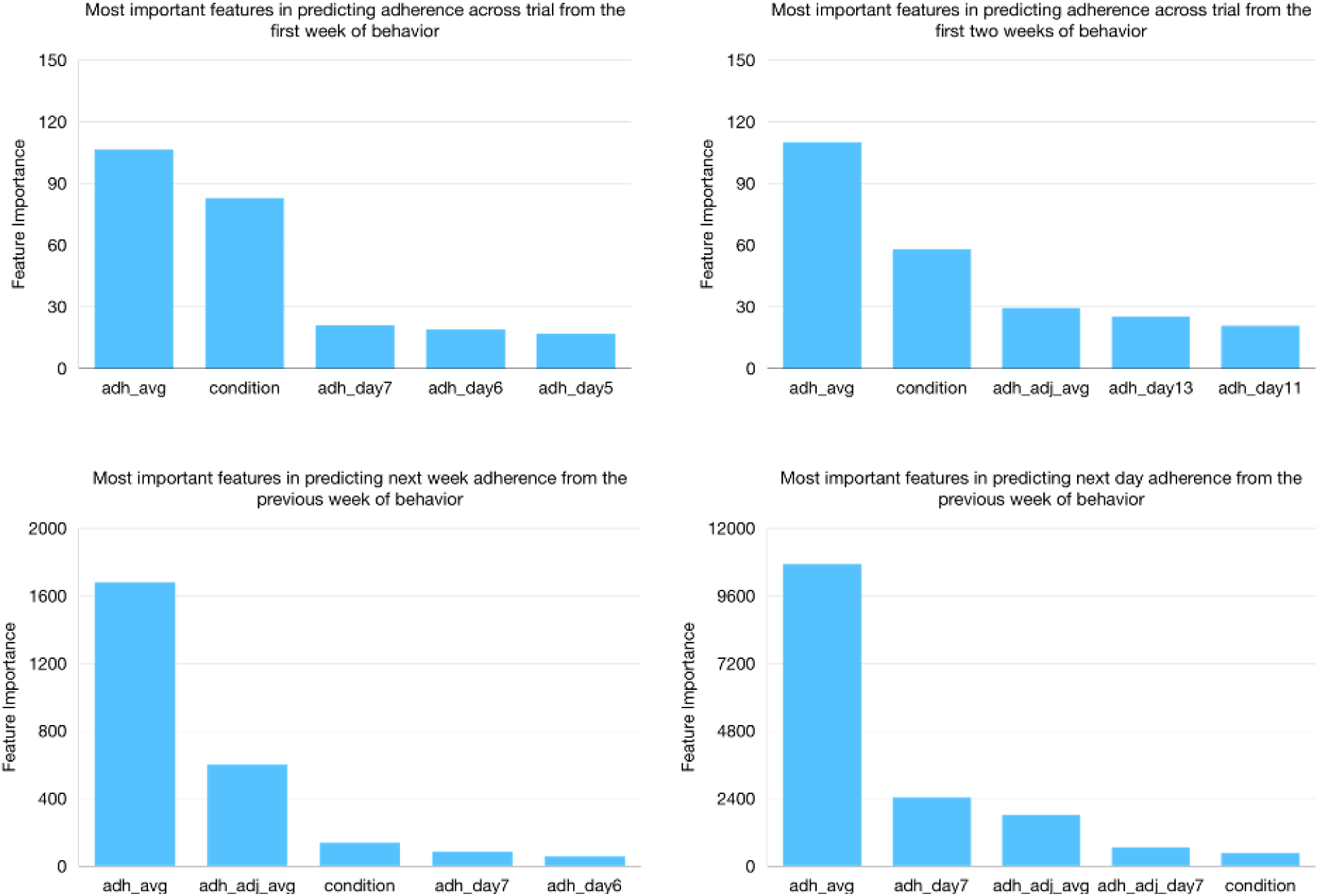
Highest ranking features in importance of the four adherence model types.

## Discussion

Here, we observed that empirical measurement of medication dosing can provide a large source of data for prediction of medication adherence, which can be subsequently achieved with high accuracy.^23^ An additional week of data significantly decreased the false positive rate. This indicates that it may be recommended, both in the context of clinical trials and in traditional clinical care, to follow patients over two weeks. Additionally, we find that clinical condition provides predictive information indicating that different conditions contribute distinctly to the probability of treatment adherence.

Results further demonstrate that as predictions become more dynamic (prediction of next week, next day), real-time measurement of dosing increases in importance and static features (e.g. condition) decrease in importance. Also, predictive accuracy increases and false positive/negative rates decrease when making dynamic predictions. The decreased relevance of condition in predicting adherence indicates that empirical measures of dosing greatly outweigh a particular type of disorder or illness such as infectious disease or psychiatric disorder.

The current study presented with limitations that affect the generalizability of the derived models. Specifically, the data is primarily collected in the context of clinical trial research. Results may not generalize to real-world samples and models should be updated with real-world data. Further, the data collected is biased towards patients with mental health disorders. Model generalizability will require integration of data from other disorder subtypes. Finally, the data was exclusively collected using the AiCure platform. Rates of adherence may be affected by the tolerability or other confounds associated with the technology used to assess adherence. This interaction between technology and behavior affects the generalizability of the results requiring further replication and integration of data from other platforms.

In sum, results indicate that new technologies that directly monitor medication dosing can be utilized to build dynamic predictive models of adherence to treatment. These predictions have both research and clinical applications that ultimately can be utilized to greatly improve treatment engagement across clinical indications.

## Data Availability

Data was collected through the use of AiCure software platform and is only available for internal use.

## References

1. Lee H, Park JH, Floyd JS, Park S, Kim HC. Combined Effect of Income and Medication Adherence on Mortality in Newly Treated Hypertension: Nationwide Study of 16 Million Person-Years. J Am Heart Assoc Cardiovasc Cerebrovasc Dis. 2019;8(16). doi:10.1161/JAHA.119.013148

2. Xie Z, St. Clair P, Goldman DP, Joyce G. Racial and ethnic disparities in medication adherence among privately insured patients in the United States. PLoS ONE. 2019;14(2). doi:10.1371/journal.pone.0212117

3. Hanina A, Kessler G. Apparatus and method for recognition of patient activities when obtaining protocol adherence data. Published online March 22, 2016. Accessed April 6, 2020. https://patents.google.com/patent/US9293060/en?oq=hanina+and+kessler++No+9%2c293%2c060

4. Hafezi H, Robertson TL, Moon GD, Au-Yeung K-Y, Zdeblick MJ, Savage GM. An Ingestible Sensor for Measuring Medication Adherence. IEEE Trans Biomed Eng. 2015;62(1):99–109. doi:10.1109/TBME.2014.2341272

5. Labovitz DL, Shafner L, Reyes Gil M, Virmani D, Hanina A. Using Artificial Intelligence to Reduce the Risk of Nonadherence in Patients on Anticoagulation Therapy. Stroke. 2017;48(5):1416–1419. doi:10.1161/STROKEAHA.116.016281

6. Sim I. Mobile Devices and Health. N Engl J Med. 2019;381(10):956–968. doi:10.1056/NEJMra1806949

7. Barfod T, Sorensen H, Nielsen H, Rodkjaer L, Obel N. “Simply forgot” is the most frequently stated reason for missed doses of HAART irrespective of degree of adherence. HIV Med. 2006;7(5):285–290. doi:10.1111/j.1468-1293.2006.00387.x

8. Hughes RG, Ortiz E. Medication Errors: Why they happen, and how they can be prevented. AJN Am J Nurs. 2005;105(Supplement):14–24. doi:10.1097/00000446-200503001-00005

9. Yamada K, Watanabe K, Nemoto N, et al. Prediction of medication noncompliance in outpatients with schizophrenia: 2-year follow-up study. Psychiatry Res. 2006;141(1):61–69. doi:10.1016/j.psychres.2004.07.014

10. DiMatteo MR, Lepper HS, Croghan TW. Depression is a risk factor for noncompliance with medical treatment: meta-analysis of the effects of anxiety and depression on patient adherence. Arch Intern Med. 2000;160(14):2101–2107. doi:10.1001/archinte.160.14.2101

11. Martin LR, Williams SL, Haskard KB, Dimatteo MR. The challenge of patient adherence. Ther Clin Risk Manag. 2005;1(3):189–199.

12. Ascher-Svanum H, Zhu B, Faries DE, et al. The cost of relapse and the predictors of relapse in the treatment of schizophrenia. BMC Psychiatry. 2010;10:2. doi:10.1186/1471-244X-10-2

13. Rosenheck R, Leslie D, Sint K, et al. Cost-Effectiveness of Comprehensive, Integrated Care for First Episode Psychosis in the NIMH RAISE Early Treatment Program. Schizophr Bull. 2016;42(4):896–906. doi:10.1093/schbul/sbv224

14. Martani A, Geneviève LD, Poppe C, Casonato C, Wangmo T. Digital pills: a scoping review of the empirical literature and analysis of the ethical aspects. BMC Med Ethics. 2020;21(1):3. doi:10.1186/s12910-019-0443-1

15. Story A, Aldridge RW, Smith CM, et al. Smartphone-enabled video-observed versus directly observed treatment for tuberculosis: a multicentre, analyst-blinded, randomised, controlled superiority trial. The Lancet. 2019;393(10177):1216–1224. doi:10.1016/S0140-6736(18)32993-3

16. Brown MT, Bussell JK. Medication Adherence: WHO Cares? Mayo Clin Proc. 2011;86(4):304–314. doi:10.4065/mcp.2010.0575

17. Haynes RB, Taylor DW, Sackett DL, Gibson ES, Bernholz CD, Mukherjee J. Can simple clinical measurements detect patient noncompliance? Hypertens Dallas Tex 1979. 1980;2(6):757–764. doi:10.1161/01.hyp.2.6.757

18. Faries DE, Heiligenstein JH, Tollefson GD, Potter WZ. The double-blind variable placebo lead-in period: results from two antidepressant clinical trials. J Clin Psychopharmacol. 2001;21(6):561–568. doi:10.1097/00004714-200112000-00004

19. Bain EE, Shafner L, Walling DP, et al. Use of a Novel Artificial Intelligence Platform on Mobile Devices to Assess Dosing Compliance in a Phase 2 Clinical Trial in Subjects With Schizophrenia. JMIR MHealth UHealth. 2017;5(2):e18. doi:10.2196/mhealth.7030

20. Pafka S. Szilard/Benchm-Ml.; 2020. Accessed April 30, 2020. https://github.com/szilard/benchm-ml

21. Natekin A, Knoll A. Gradient boosting machines, a tutorial. Front Neurorobotics. 2013;7. doi:10.3389/fnbot.2013.00021

22. Chen T, Guestrin C. XGBoost: A Scalable Tree Boosting System. Proc 22nd ACM SIGKDD Int Conf Knowl Discov Data Min - KDD 16. Published online 2016:785–794. doi:10.1145/2939672.2939785

23. Lauffenburger JC, Franklin JM, Krumme AA, et al. Predicting Adherence to Chronic Disease Medications in Patients with Long-term Initial Medication Fills Using Indicators of Clinical Events and Health Behaviors. J Manag Care Spec Pharm. 2018;24(5):469–477. doi:10.18553/jmcp.2018.24.5.469

